# Evaluating Enhanced LLMs for Precise Mental Health Diagnosis from Clinical Notes

**DOI:** 10.1101/2024.12.16.24317648

**Authors:** Lokesh Boggavarapu, Vineet Srivastava, Amit Maheswar Varanasi, Yingda Lu, Runa Bhaumik

**Affiliations:** University of Illinois Chicago

**Keywords:** Large Language Model, Retrieval Augmented Generation, Mental Health, Clinical Notes

## Abstract

Anxiety, depression, and other mental health conditions are affecting millions of people worldwide each year. However, limited access to mental health professionals and the stigma surrounding mental illness often deter individuals from seeking help. Many areas, especially rural and underserved communities, face a significant shortage of mental health professionals, making it difficult for individuals to access timely support and treatment. Traditional therapy can be expensive, time-consuming, and intimidating, discouraging individuals from seeking care and delaying essential treatment. The goal of this project is to harness the power of large language models

In the medical domain, large language models (LLMs) have the potential to significantly enhance clinical practice by assisting with tasks such as diagnostic support, therapeutic interventions, and summarization. However, these models often generate inaccurate responses, or “hallucinations,” when faced with queries they cannot effectively handle, raising concerns in the medical community. To address the limitations of LLMs, Retrieval-Augmented Generation (RAG) was leveraged to enhance their performance. By integrating external knowledge sources such as ICD-10-CM guidelines and psychiatric diagnostic manuals, RAG enables LLMs to retrieve relevant information in real time to support their predictions. This study examines whether LLMs can understand and accurately predict mental health-related medical codes from clinical notes. These codes are crucial for clinical documentation and treatment planning. We tested several LLMs (e.g., GPT, LLaMA, Gemini-Pro) enhanced with reliable resources like ICD-10-CM guidelines to evaluate their ability to identify and understand mental health terms and ICD-10-CM codes in psychiatric clinical notes. Our findings reveal that current models lack a robust understanding of the meaning and nuances of these codes, limiting their reliability for mental health applications. This underscores the need for improved strategies to represent and integrate these complex alphanumeric codes within LLMs. Enhancing their capability to accurately process mental health terminologies would make LLMs more reliable and trustworthy tools for mental health professionals, ultimately supporting better care and outcomes for patients.

## Introduction

Mental health is a global concern in public health today. According to the US National Institute of Mental Health (NIMH)^1^, more than one in five U.S. adults live with a mental illness (59.3 million in 2022; 23.1% of the U.S. adult population. A large part of managing mental health involves natural language, including the assessment of symptoms and signs of mental health disorders and interventions such as talk therapies^2^ The textual and conversational data from these interactions offer a valuable resource for improving research and practical approaches to mental health care. One way to prevent serious mental conditions is to identify the symptoms in earlier stages. Natural Language Processing (NLP), a field of computer science focused on enabling computers to process and interpret unstructured natural language data, has shown great potential in supporting mental health-related tasks. These applications include identifying specific mental health conditions^3^, developing emotion-support chatbots^4^, and assisting with interventions^5^. NLP leverages data from diverse sources, such as clinical records^6^ and social media platforms^7,8^. In mental healthcare, Large Language Models (LLMs), a recent breakthrough in NLP, have opened new possibilities for innovative mental healthcare^9,10^. LLMs have the potential to streamline processes by quickly analyzing patient data, summarizing therapy sessions, and assisting with complex diagnostic challenges, saving valuable time for both users and healthcare providers^11^. In recent studies, Psy-LLM^12^, Mental-LLM^13^, MentalLLaMA^14^, and MindWatch^15^, were used to generate predictions or counselling based on input prompts related to mental health conditions. This signifies a growing interest in leveraging generative LLMs and prompt learning for mental health-related tasks.

Clinical notes in the psychiatry domain contain a wide range of information that helps mental health professionals assess, diagnose, treat, and monitor patients with mental health conditions. These notes serve as a detailed record of patient encounters, providing essential insights into the patient’s mental state, history, and treatment progress. LLMs have a high potential to analyze psych clinical notes and identify diagnoses in an automated way. However, studies on LLMs for diagnosing psychiatric disorders via automatic coding remain limited. To the best of our knowledge, this is the first study that evaluates LLMs as an automated ICD code detection method using clinical notes from the psychiatry domain.

ICD coding often faces errors and inconsistencies due to variations in how coders interpret and apply the coding rules and criteria or the omission of relevant codes. To tackle these challenges, automatic ICD coding has gained prominence as a research focus in clinical medicine, utilizing machine learning and NLP algorithms^20,21^. Large language models (LLMs), such as GPT models, Llama, and Gemini, have been also explored for this task^22,23^. However, these studies highlight several challenges, including LLMs’ limited domain-specific knowledge and vocabulary, the complexity of multi-label and long-tail code spaces, and vulnerability to noisy or adversarial inputs^22,24^

This study evaluates the baseline coding performance of various LLMs, including GPT-4, Gemini-1.5-Flash, and LLaMa-70b Chat. It also investigates the effectiveness of different LLM configurations, including retrieval-augmented generation (RAG)^25^ and zero-shot prompting in detecting ICD-10 codes in the context of real-world clinical narratives or notes from the EHR. Using a systematic approach, the models were instructed to generate medical codes based on their descriptions. This assessment aims to identify areas for improvement in code-mapping accuracy and provide insights for future advancements.

## II Methodology

### A Data Preparation

We extracted Clinical Modification (ICD-10-CM) codes from the University of Illinois health system for mental health disorders and diseases. We have randomly chosen 50 clinical notes for our experiment. All sensitive information was removed to maintain privacy. Our data was stored on secure HIPAA-compliant servers. We used the GPT-4o model provided by Microsoft’s Azure OpenAI service (www.portal.azure.com), Gemini-1.5 Flash within a secured Google cloud environment and Llama3.2 in a locally secured server to generate AI-based predictions (the probability) for the diagnoses of mental health disorders. Clinical diagnoses made by qualified psychiatrists were treated as gold standards (target) for evaluation.

### B Model Configurations

We evaluated several LLM configurations to compare their performance on ICD-10 code detection tasks:

1. **Retrieval-Augmented Generation (RAG)**^25^: It is a framework that combines the strengths of two core components in Natural Language Processing (NLP): retrieval and generation. In the context of psychiatry, where specialized and accurate knowledge is crucial, RAG leverages external knowledge sources like ICD-10-CM guidelines to generate contextually relevant and highly informed responses.
2. **Zero-Shot**^26^: Models were applied without prior training on ICD-10-specific tasks, relying on general language capabilities.

### C Models

The following LLMs were selected based on their prominence and applicability:

#### LLAMA 3.2^27^

Developed by Meta, Llama 3.2 is a collection of multilingual large language models designed to process both text and images. It includes models with 1 billion and 3 billion parameters optimized for on-device applications, as well as vision models with 11 billion and 90 billion parameters capable of handling multimodal inputs. This versatility makes Llama 3.2 suitable for various applications, from augmented reality to document analysis.

#### Gemini-1.5-Flash^28^

Released by Google DeepMind, Gemini 1.5 Flash is a lightweight, multimodal model optimized for speed and efficiency. It can process inputs such as audio, images, video, and text, generating text responses suitable for tasks like code generation, data extraction, and text editing. The model is designed for high-frequency tasks where performance and cost-efficiency are paramount

#### GPT-4o^29^

Developed by OpenAI, GPT-4 is a large multimodal model capable of processing both text and image inputs to generate text outputs. It excels in tasks requiring advanced reasoning, complex instruction, understanding, and creativity. GPT-4 has been integrated into various applications, including chatbots and content generation tools, demonstrating its versatility and robustness.

### D Evaluation Metrics

Performance was measured using the following metrics:

#### Jaccard Similarity

Measures overlap between predicted and actual ICD-10 codes. The Jaccard coefficient is calculated by dividing the size of the intersection of two sets by the size of their union:

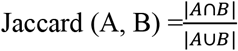

#### Macro Precision^30^

It is the proportion of true positive results (correct predictions) out of all the results that were predicted as positive. In this work, for multilabel classification, the precision was calculated independently for each label and then averaged across labels by treating all labels equally.

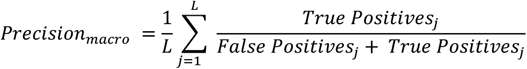

#### Macro Recall^30^

Recall is calculated independently for each label and then averaged across labels.

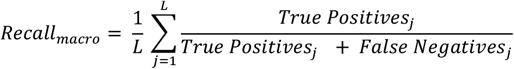

where L is the total number of labels. This approach treats each label equally, regardless of its frequency in the dataset.

#### F1 Score^30^

The harmonic mean of Precision and Recall offers a balanced metric.

#### Hamming Loss^30^

Reflects the fraction of labels that are incorrectly predicted.

#### Completely Different

Refers to instances where the predicted set of labels has no overlap with the actual set of labels.

## III Results

### Zero-Shot Configuration

GPT-4o emerged as the top-performing model, achieving the highest scores across all key metrics, including Jaccard Similarity (0.674), Precision (0.793), Recall (0.717), and F1 Score (0.730), with the lowest Hamming Loss (0.032) and only 12 completely different outputs. This indicates that GPT-4o is robust even without additional context or task-specific training. Google demonstrated moderate performance, achieving Jaccard Similarity (0.573) and F1 Score (0.612), though it struggled with 26 completely different outputs and higher Hamming Loss (0.042) than GPT-4o. LLaMA showed the weakest performance, with the lowest scores for Jaccard Similarity (0.509), Precision (0.559), and F1 Score (0.536). Additionally, it produced the highest number of completely different outputs (39) and exhibited a higher Hamming Loss (0.043) compared to GPT-4o and Google.

**Table 1.**
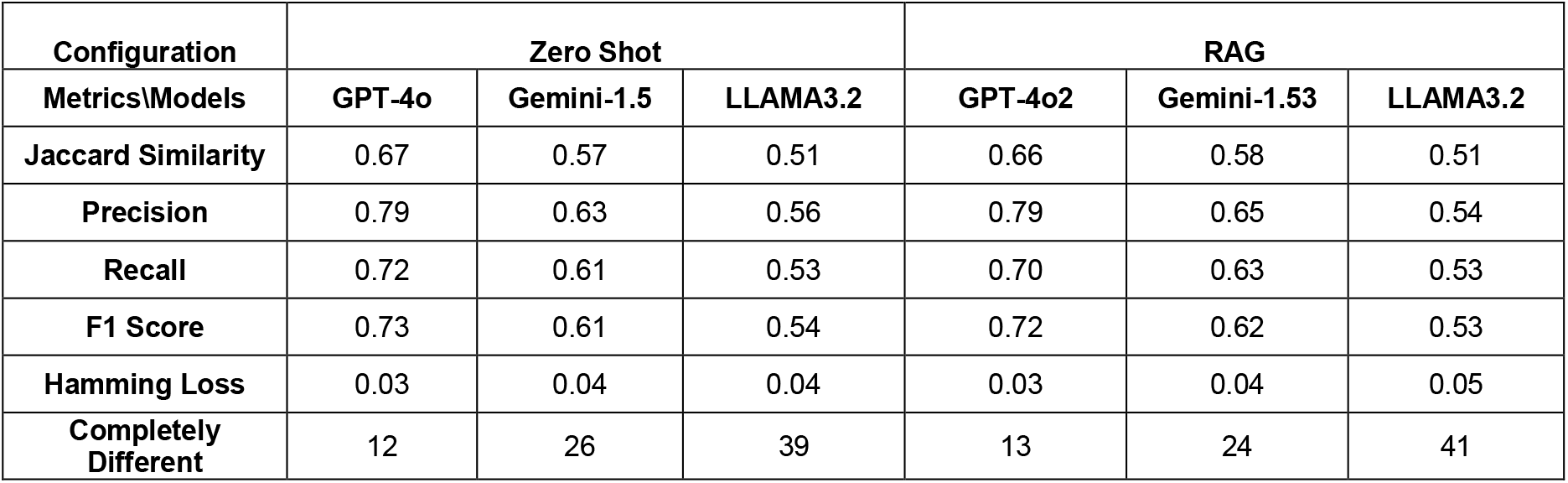
Comparisons of different metrics for different models in two configurations.

### RAG Configuration

GPT-4o maintained its lead under the RAG setup, with metrics showing slight improvements or stability: Jaccard Similarity (0.664), Precision (0.786), Recall (0.705), F1 Score (0.720), and Hamming Loss (0.032). The model also reduced its number of completely different outputs to 13, highlighting its effectiveness in leveraging additional context. Google exhibited noticeable improvement in the RAG configuration, with increased Jaccard Similarity (0.578), Precision (0.646), and F1 Score (0.624). However, it still produced 24 completely different outputs, reflecting some inconsistency in predictions. LLaMA showed limited improvement in the RAG configuration, with Jaccard Similarity (0.506), Precision (0.544), and F1 Score (0.531) remaining low. It also had the highest Hamming Loss (0.047) and the most completely different outputs (41), suggesting minimal benefit from the additional context provided by RAG. The model also reduced its number of completely different outputs to 13, highlighting its effectiveness in leveraging additional context.

#### Detection of the higher level of code

Clinical notes might not always contain enough detail to assign the full ICD-10 code accurately. Focusing on three digits ensures the evaluation reflects what the notes actually provide, avoiding over-reliance on assumptions. We have evaluated three models with zero-shot prompting techniques for a general category of diseases or conditions. Comparison results with fine-grained evaluation are shown in Table 2.

**Table 2.** Comparisons of different metrics for different models in two configurations.

| Configuration<br>Metrics\Models | Zero-Shot |  |  | Zero-Shot (Higher Level) |  |  |
| --- | --- | --- | --- | --- | --- | --- |
|  | GPT-4o | Gemini-1.5 Flash | LLAMA3.2 | GPT-4o | Gemini-1.5 Flash | LLAMA3.2 |
| Jaccard Similarity | 0.67 | 0.57 | 0.51 | 0.77 | 0.69 | 0.61 |
| Precision | 0.79 | 0.63 | 0.56 | 0.87 | 0.76 | 0.7 |
| Recall | 0.72 | 0.61 | 0.53 | 0.84 | 0.75 | 0.66 |
| F1 Score | 0.73 | 0.61 | 0.54 | 0.83 | 0.74 | 0.66 |
| Hamming Loss | 0.03 | 0.04 | 0.04 | 0.04 | 0.05 | 0.06 |
| Completely Different | 12 | 26 | 39 | 4 | 15 | 21 |

For a higher level of code detection, GPT-4o achieved the highest Jaccard Similarity (0.77), Precision (0.87), Recall (0.84), and F1 Score (0.83). It also showed the lowest number of Completely Different predictions (4). These results demonstrated the strongest ability to predict ICD codes, particularly in higher-level evaluations.

#### Human Evaluation

The evaluation of the model (GPT4o) against human-coded ICD diagnoses in mental health clinical notes revealed several discrepancies. While the GPT-4o model demonstrated the ability to pick up relevant codes based on text patterns and keywords in the assessment/plan, it frequently missed certain codes identified by clinicians. For example, the model often failed to capture multiple codes that clinicians identified, including recurrent depressive disorders (e.g., F33.2, F33.41), anxiety disorders (e.g., F41.9, F41.1), and substance use disorders (e.g., F11.90, F12.90, F10.21).In many cases, the model assigned the primary code correctly but failed to include additional codes that the doctor deemed relevant, potentially missing conditions. The model occasionally assigns codes for conditions that are part of the family history/initial diagnosis which the doctor disregarded at a later stage(e.g. F43.9 F41.1 F10.10 F17.200) from which the doctor only mentioned F41.1 Also, the model occasionally assigned incorrect codes based on partial matches in text, such as assigning F41.1 (Generalized Anxiety Disorder) for mentions of anxiety, even when the clinician specified a more precise diagnosis (e.g., F43.23 (Adjustment Disorder with Mixed Anxiety and Depressed Mood)).

## IV Discussion

### A Analysis of Results

We evaluated the performance of various Large Language Models (LLMs), including GPT-4o, Google, and LLaMA, for ICD coding tasks under two configurations: Zero-Shot and Retrieval-Augmented Generation (RAG). The evaluation highlights significant differences in the models’ capabilities for ICD coding tasks: GPT-4o consistently outperformed Google and LLaMA in both configurations, demonstrating superior precision, recall, and F1 scores, as well as the lowest Hamming Loss. Its ability to maintain high performance under both Zero-Shot and RAG conditions underscores its robustness and adaptability. **Google** displayed moderate performance, benefiting from the RAG setup but still falling short of GPT-4o’s reliability and consistency. LLaMA lagged in all metrics, with minimal gains from RAG, suggesting that it is not well-suited for medical coding tasks in its current state. The RAG configuration improved the performance of all models to varying degrees, indicating that integrating external knowledge sources can enhance LLMs’ ability to handle complex and domain-specific tasks. In addition, GPT-4o consistently outperformed Google and LLaMA across all metrics in both zero-shot and higher-level evaluations.

### B Limitations

Due to the sensitive nature of the information and the cost associated with the computing environment, we limited our evaluation to 50 notes, which may impact the comprehensiveness of the assessment. In the RAG approach, we have only used one guideline, ICD-10-CM, which may have a limited impact on retrieval.

### C Implications for Future Work

Future research should explore further customization of LLMs with domain-specific training data, particularly in psychiatry. Other reliable documents, such as APA guidelines, UMLS code mapping may improve the detection accuracy.

## V Conclusion

This study demonstrates the potential of large language models for detecting ICD-10 codes in psychiatry notes, with GPT-4o (RAG) showing the highest overall performance across most metrics. The findings indicate that LLMs, especially when combined with retrieval-augmented techniques, could support automated ICD-10 coding in clinical practice. However, achieving exact matches remains challenging, and further refinement of multi-agent and RAG methods could lead to even greater accuracy. This work underscores the transformative potential of LLMs in healthcare, particularly for psychiatry applications where language complexity is high.

## Data Availability

All data produced in the present study are available upon reasonable request to the authors

## VI. Acknowledgements

The authors would like to thank two medical coder experts from the Psychiatry department at the University of Illinois Chicago for their invaluable feedback.

## VII Ethical Considerations

All personal and sensitive information within the dataset has been anonymized to safeguard individuals’ identities and privacy. Data is stored securely and accessed only by authorized personnel. This study has been approved as exempt under the University of Illinois Chicago’s Institutional Review Board (IRB) protocol.

## References

1. NIMH. Of Mental Health (NIMH), N. I. Mental Illness (2023).

2. Sadock, B. J. et al. Kaplan & Sadock’s Synopsis of Psychiatry: Behavioral Sciences/Clinical Psychiatry. Vol 2015. Wolters Kluwer Philadelphia, PA

3. Zhang T, Schoene AM, Ji S, Ananiadou S. Natural language processing applied to mental illness detection: a narrative review. npj Digit Med. 2022;5(1):46. doi:10.1038/s41746-022-00589-7

4. Malgaroli M, Hull TD, Zech JM, Althoff T. Natural language processing for mental health interventions: a systematic review and research framework. Transl Psychiatry. 2023;13(1):309. doi:10.1038/s41398-023-02592-2

5. Li H, Zhang R, Lee YC, Kraut RE, Mohr DC. Systematic review and meta-analysis of AI-based conversational agents for promoting mental health and well-being. npj Digit Med. 2023;6(1):236. doi:10.1038/s41746-023-00979-5

6. Irving, J. et al. Using natural language processing on electronic health records to enhance detection and prediction of psychosis risk. Published online 2021.

7. Li M, Hua Y, Liao Y, et al. Tracking the Impact of COVID-19 and Lockdown Policies on Public Mental Health Using Social Media: Infoveillance Study. J Med Internet Res. 2022;24(10):e39676. doi:10.2196/39676

8. Skaik R, Inkpen D. Using Social Media for Mental Health Surveillance: A Review. ACM Comput Surv. 2021;53(6):1–31. doi:10.1145/3422824

9. Ji S, Zhang T, Yang K, Ananiadou S, Cambria E. Rethinking Large Language Models in Mental Health Applications. Published online 2023. doi:10.48550/ARXIV.2311.11267

10. Van Heerden AC, Pozuelo JR, Kohrt BA. Global Mental Health Services and the Impact of Artificial Intelligence–Powered Large Language Models. JAMA Psychiatry. 2023;80(7):662. doi:10.1001/jamapsychiatry.2023.1253

11. Obradovich N, Khalsa SS, Khan WU, et al. Opportunities and risks of large language models in psychiatry. NPP—Digit Psychiatry Neurosci. 2024;2(1):8. doi:10.1038/s44277-024-00010-z

12. Lai T, Shi Y, Du Z, et al. Psy-LLM: Scaling up Global Mental Health Psychological Services with AI-based Large Language Models. Published online 2023. doi:10.48550/ARXIV.2307.11991

13. Xu X, Yao B, Dong Y, et al. Mental-LLM: Leveraging Large Language Models for Mental Health Prediction via Online Text Data. Proc ACM Interact Mob Wearable Ubiquitous Technol. 2024;8(1):1–32. doi:10.1145/3643540

14. Yang K, Zhang T, Kuang Z, Xie Q, Huang J, Ananiadou S. MentaLLaMA: Interpretable Mental Health Analysis on Social Media with Large Language Models. In: Proceedings of the ACM Web Conference 2024. ACM; 2024:4489–4500. doi:10.1145/3589334.3648137

15. Bhaumik R, Srivastava V, Jalali A, Ghosh S, Chandrasekharan R. MindWatch: A Smart Cloud-based AI solution for Suicide Ideation Detection leveraging Large Language Models. Published online September 26, 2023. doi:10.1101/2023.09.25.23296062

16. WHO. World Health Organization. History of the Development of the ICD. (https://cdn.who.int/media/docs/default-source/classification/icd/historyoficd.pdf)

17. who. International Statistical Classification of Diseases and Related Health Problems 10th Revision. https://icd.who.int/browse10/2019/en

18. World Health Organization. Importance of ICD. 2024. https://www.who.int/standards/classifications/frequently-asked-questions/importance-of-icd

19. Hirsch JA, Nicola G, McGinty G, et al. ICD-10: History and Context. AJNR Am J Neuroradiol. 2016;37(4):596–599. doi:10.3174/ajnr.A4696

20. Dong H, Falis M, Whiteley W, et al. Automated clinical coding: what, why, and where we are? npj Digit Med. 2022;5(1):159. doi:10.1038/s41746-022-00705-7

21. Ponthongmak W, Thammasudjarit R, McKay GJ, Attia J, Theera-Ampornpunt N, Thakkinstian A. Development and external validation of automated ICD-10 coding from discharge summaries using deep learning approaches. Informatics in Medicine Unlocked. 2023;38:101227. doi:10.1016/j.imu.2023.101227

22. Soroush A, Glicksberg BS, Zimlichman E, et al. Large Language Models Are Poor Medical Coders — Benchmarking of Medical Code Querying. NEJM AI. 2024;1(5). doi:10.1056/AIdbp2300040

23. Lahat A, Sharif K, Zoabi N, et al. Assessing Generative Pretrained Transformers (GPT) in Clinical Decision-Making: Comparative Analysis of GPT-3.5 and GPT-4. J Med Internet Res. 2024;26:e54571. doi:10.2196/54571

24. Lee SA, Lindsey T. Can Large Language Models abstract Medical Coded Language? Published online June 6, 2024. Accessed November 17, 2024. http://arxiv.org/abs/2403.10822

25. Lewis P, Perez E, Piktus A, et al. Retrieval-Augmented Generation for Knowledge-Intensive NLP Tasks. Published online 2020. doi:10.48550/ARXIV.2005.11401

26. Brown TB, Mann B, Ryder N, et al. Language Models are Few-Shot Learners. Published online 2020. doi:10.48550/ARXIV.2005.14165

27. Meta. Llama 3.2: Revolutionizing edge AI and vision with open, customizable models. 2024. https://ai.meta.com/blog/llama-3-2-connect-2024-vision-edge-mobile-devices/

28. Google. Gemini’s big upgrade: Faster responses with 1.5 Flash, expanded access and more.

29. OpenAI. We’re Announcing GPT-4o, Our New Flagship Model That Can Reason across Audio, Vision, and Text in Real Time.; 2024.

30. Tsoumakas G, Katakis I. Multi-Label Classification: An Overview. International Journal of Data Warehousing and Mining. 2007;3(3):1–13. doi:10.4018/jdwm.2007070101

